# First-Line Opioids and Short-Term All-Cause Emergency Department Return After Headache Visits: A Two-Center Comparative Cohort Study

**DOI:** 10.64898/2026.07.16.26358169

**Authors:** Alon Gorenshtein, Yosef Adiniaev, Tom Liba, Eyal Klang, Oved Daniel

## Abstract

**Objective:** To compare first-line emergency department (ED) treatment classes for acute headache on short-term all-cause ED return and index admission across two independent health systems.

**Background:** ED trials of acute headache treatment are judged on in-ED pain relief, a documented endpoint that is recorded incompletely and shifts with the scoring rule, and is a weak surrogate for what happens after discharge. All-cause ED return after an index headache visit (any subsequent ED encounter within the window) has not been used to compare first-line treatments at scale, and society guidance favors dopamine-receptor antagonists while recommending against routine opioids.

**Methods:** Retrospective two-center cohort of adults treated for headache in the ED, using MIMIC-IV-ED (Beth Israel Deaconess Medical Center, 2011-2019) and MC-MED (Stanford, 2020-2022). The first-line class was the earliest qualifying acute agent. The primary contrast was opioids versus dopamine-receptor antagonists (the guideline-preferred class). Outcomes were 72-hour and 7-day all-cause ED return (among discharged patients; any subsequent ED encounter within the window) and index hospital admission. Confounding by indication was addressed with propensity overlap weighting; associations are reported as adjusted risk ratios (RRs) with bootstrap 95% CIs and E-values. Estimates were pooled with a site term and examined per site.

**Results:** Among 13,285 treated adults (10,799 MIMIC-IV-ED; 2,486 MC-MED), opioid recipients were older and higher-acuity than dopamine-antagonist recipients (index admission 38.1% vs 16.4%). In the MIMIC-IV-ED discharged primary-contrast population, overlap weighting reduced the maximum standardized mean difference from 0.35 to 0.002; pooled and site-specific balance diagnostics are provided in the Supplement. First-line opioids remained associated with a higher 72-hour all-cause ED return (6.8% vs 3.8%; adjusted RR 1.79; 95% CI 1.31 to 2.33), 7-day return (10.7% vs 6.6%; RR 1.62; 95% CI 1.28 to 1.98), and index admission (RR 2.32; 95% CI 2.11 to 2.58, consistent with strong residual severity differences in patients selected for opioids). The direction of association was concordant across both health systems, although MC-MED return estimates were imprecise given the smaller opioid-treated discharged sample. In MIMIC-IV-ED, the cumulative all-cause return incidence by treatment class separated by day 3 and persisted through 30 days. The direction was consistent, though attenuated and no longer statistically significant, when the outcome was restricted to a headache-specific return (72-hour RR 1.31; 95% CI 0.91 to 1.88); the direction persisted for the composite of admission or 72-hour return, which does not condition on discharge but is influenced by the more confounded admission component (RR 2.16; 95% CI 1.98 to 2.39).

**Conclusion:** Across two health systems, first-line opioid treatment for ED headache was associated with higher all-cause short-term ED return among discharged patients and higher index admission than dopamine antagonists. These observational associations reflect downstream all-cause ED utilization after an index headache visit rather than confirmed headache recurrence or treatment failure; they are consistent with guideline-concordant, opioid-sparing first-line treatment and warrant prospective confirmation.

**Plain Language Summary:** Emergency departments treat headaches with several different medicines, but the usual way of judging which works, the pain score recorded during the visit, is often missing or inconsistent. Using two large hospital systems and a clearer outcome, whether patients came back to the emergency department for any reason, we found that patients first treated with opioids returned within 72 hours about 1.8 times as often as those given the guideline-preferred dopamine-blocking medicines and were admitted more than twice as often. These patterns pointed the same direction in both hospital systems after adjustment for the measured differences available in both databases. Because this was an observational comparison and returns were counted for any reason, the findings are consistent with using guideline-preferred non-opioid medicines first, rather than proof that opioids worsen headache.

**Graphical abstract:** Standalone visual summary of the two-center comparison: among adults treated for headache in the emergency department, first-line opioids were associated with a higher all-cause ED return and higher index admission than guideline-preferred dopamine antagonists, whereas the headache-specific return was attenuated and crossed the line of no difference. Index admission is shown as a secondary acute-care outcome and should be interpreted cautiously because it is highly susceptible to confounding by baseline severity.

## Introduction

Acute headache is among the most common reasons adults present to the emergency department (ED), and first-line treatment varies widely between dopamine-receptor antagonists, nonsteroidal anti-inflammatory drugs, triptans, intravenous magnesium, and opioids.[1,2] Randomized trials and society guidance favor parenteral dopamine antagonists such as metoclopramide and prochlorperazine and recommend against routine opioids, which are less effective for migraine and carry dependence and medication-overuse risk.[3,4] Yet opioids remain a frequent first-line choice in routine ED care.[5,6]

The evidence base that shapes this guidance is anchored to an in-ED pain-relief endpoint measured at fixed times in trials. In routine care, that documented relief is recorded incompletely and shifts materially with the reassessment window and the score-selection rule, so it is a weak surrogate for what happens after the patient leaves.[7,8] The outcome that matters to patients and to health systems is whether they return to the ED after the index visit, whatever the reason. Short-term all-cause ED return and index admission are objective, recorded without a protocol, and directly relevant to ED quality, but they have not been used to compare first-line headache treatments at scale, and the few revisit analyses that exist are single-center and cannot separate a treatment signal from generalizable practice.[9]

We used two independent ED databases from different health systems and electronic health records to compare first-line acute treatment classes for adult headache on the short-term return visit and index admission. We framed the comparison as an active-comparator analysis of opioids versus guideline-preferred dopamine antagonists, addressed confounding by indication with propensity overlap weighting, and examined whether any association was directionally concordant across both sites. Because treatment choice is not randomized, we report risk-adjusted associations and quantify their sensitivity to unmeasured confounding rather than claiming a causal effect.

## Methods

### Study Design and Setting

This was a retrospective, two-center comparative cohort study structured as an active-comparator, new-user analysis. The primary cohort was MIMIC-IV-ED (version 2.2), a de-identified database of ED visits to Beth Israel Deaconess Medical Center between 2011 and 2019.[10] External comparison used MC-MED (version 1.0.0), a de-identified ED database from Stanford University Medical Center spanning approximately 2020 to 2022 on a different electronic health record.[11] Reporting followed STROBE with the RECORD extension for routinely collected data.[12] Treatment was not randomized; no causal effect was estimated, and associations are reported as adjusted.

### Cohort

Adults (age >= 18 years) with an ED visit for headache were identified by a triage chief-complaint term (headache, head ache, migraine, cephalgia, or cephalalgia) or a primary or nonspecific-headache ED diagnosis (International Classification of Diseases, Ninth Revision codes 346.x, 307.81, 339.x, or 784.0; Tenth Revision codes G43, G44, or R51; eTable 1). The treated cohort received at least one qualifying acute agent. To satisfy the new-user framing, the unit of analysis was the first treated headache visit per patient. Identical definitions were applied in both databases.

### Exposure

The first-line treatment class was the earliest qualifying acute agent by timestamp: dopamine-receptor antagonist/antiemetic agents (metoclopramide, prochlorperazine, droperidol, chlorpromazine, promethazine), opioids, nonsteroidal anti-inflammatory drugs or ketorolac, triptans or dihydroergotamine, intravenous magnesium, or acetaminophen (agent lists, eTable 2). When agents shared the earliest time, a combination containing an opioid was classified as opioid. The prespecified primary contrast was opioids versus dopamine antagonists, the guideline-preferred active comparator. A sensitivity analysis restricted the comparator to the evidence-based antimigraine agents metoclopramide, prochlorperazine, and droperidol.

### Outcomes

The primary outcome was all-cause ED return within 72 hours, defined as any subsequent ED encounter by the same patient within 72 hours of index ED departure, irrespective of the reason for return (in MIMIC-IV-ED from linked subject identifiers and disposition times; in MC-MED from the recorded interval to the next visit). The return-visit estimand is conditional on discharge from the index visit; because disposition may be associated with both treatment selection and clinical severity, these estimates are not the total effect of first-line treatment among all treated patients, and a composite of admission or 72-hour return (not conditioned on discharge) is reported as a sensitivity analysis. Secondary outcomes were the 7-day and 30-day return, index hospital admission, and the cumulative incidence of return over 30 days. Because return requires discharge, return analyses were restricted to patients discharged from the index visit; index admission was analyzed in the full treated cohort.

### Covariates

Prespecified confounders available in both databases were age, sex, race and ethnicity, triage acuity, triage pain score, mode of arrival, prior ED utilization, and time of day; the primary database additionally contributed a calendar-period term and the pooled model a site term. Data elements not available in both databases, and therefore unadjusted, included headache subtype, prior migraine history, chronic or home opioid use, pregnancy, malignancy, neuroimaging or lumbar puncture, focal neurologic deficit, and provider-level practice. Triage pain-score missingness was handled with a missing-indicator term and median substitution; triage-acuity missingness was retained as an explicit category.

### Statistical Analysis

Confounding by indication was addressed with propensity overlap weights derived from a logistic model of opioid versus dopamine-antagonist treatment on the covariates above, with standardized inputs.[13] Overlap weighting targets the population with the most clinical equipoise and yields exact mean balance on the included covariates; balance was summarized by the maximum absolute standardized mean difference. Weighted absolute risks, risk differences, and RRs were estimated for each outcome, with 95% CIs from 1000 patient-level bootstrap resamples that repeated the weighting within each replicate. Robustness to unmeasured confounding was quantified with the E-value.[14] The cumulative incidence of return over 30 days was estimated for each first-line class among discharged patients, with in-hospital death treated as a competing event. Estimates were pooled across sites with a site indicator and examined separately by site. Analyses used Python 3.9 (pandas, scikit-learn, lifelines); the random seed was fixed.

### Sensitivity analyses

Prespecified sensitivity analyses comprised (1) a headache-specific return outcome, in which the qualifying return visit was itself coded or triaged as headache; (2) restriction to migraine or primary-headache-coded index visits; (3) exclusion of index visits with a secondary-headache or red-flag diagnosis (intracranial hemorrhage, ischemic stroke or transient ischemic attack, meningitis or encephalitis, central nervous system neoplasm, head or facial trauma and intracranial injury, hydrocephalus or raised intracranial pressure, temporal arteritis, cerebral venous thrombosis, and eclampsia or pre-eclampsia); (4) a restricted dopamine comparator and an alternative medication-tie rule; and (5) a composite of index admission or 72-hour return. Covariate balance and Kish effective sample sizes are reported for every analytic population.

### Ethics

Both databases are de-identified and were accessed under credentialed PhysioNet data use agreements. The original MIMIC-IV collection was approved by the institutional review boards of the Massachusetts Institute of Technology and Beth Israel Deaconess Medical Center with a waiver of consent. This secondary analysis was non-human-subjects research and required no additional review.

## Results

### Cohort

The treated analytic cohort comprised 13,285 adults (10,799 in MIMIC-IV-ED and 2,486 in MC-MED). In the primary database, the most common first-line classes were dopamine antagonists (3,056), acetaminophen (3,361), nonsteroidal anti-inflammatory drugs (2,304), and opioids (1,895), with intravenous magnesium (167) and triptans (16) uncommon (Table 1). Baseline return rates were nearly identical across the two health systems (72-hour 4.4% and 4.1%; 7-day 7.5% and 7.6%; 30-day 15.5% and 16.4%), despite different electronic health records, regions, and calendar periods.

**Table 1.**
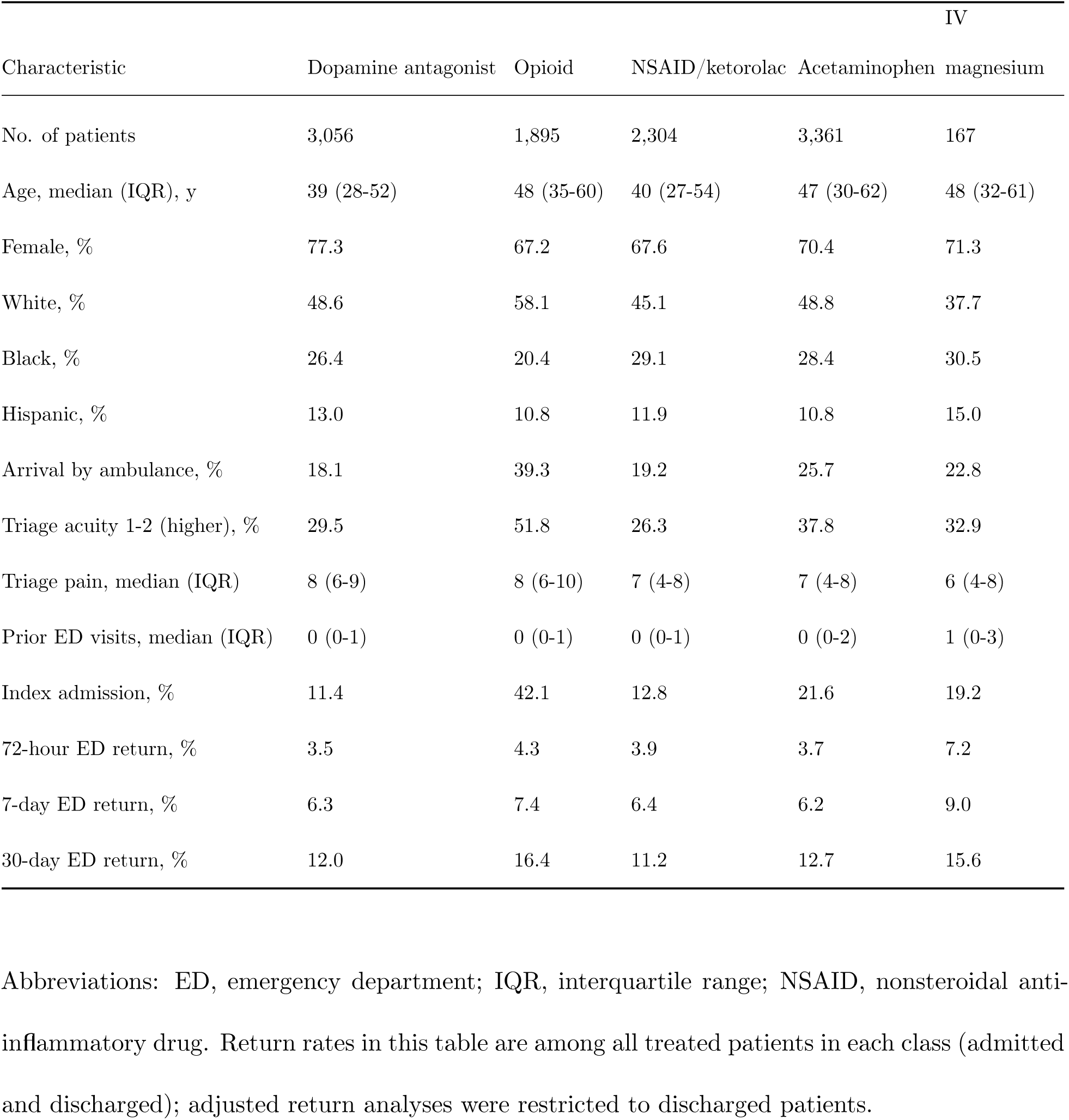
Characteristics of the treated emergency department headache cohort by first-line treatment class (MIMIC-IV-ED).

### Opioid Recipients Were Sicker at Baseline

First-line opioid recipients differed systematically from dopamine-antagonist recipients: they were older (median age 48 vs 39 years), more often arrived by ambulance (39.3% vs 18.1%), more often presented at high triage acuity (51.8% vs 29.5%), and were far more often admitted at the index visit (42.1% vs 11.4%; Table 1). This confounding by indication, in which opioids were preferentially given to higher-acuity patients, is the central threat the analysis was designed to address.

### First-Line Opioids Were Associated With More Short-Term Returns After Adjustment

In the MIMIC-IV-ED discharged primary-contrast population, overlap weighting reduced the maximum standardized mean difference from 0.35 to 0.002; pooled and site-specific balance diagnostics are provided in the Supplement; balance and effective sample sizes for every analytic population are given in eTable 9. After weighting, first-line opioids remained associated with a higher 72-hour return than dopamine antagonists among discharged patients (6.8% vs 3.8%; adjusted risk difference 2.98 percentage points, 95% CI 1.30 to 4.48; RR 1.79, 95% CI 1.31 to 2.33; Figure 3). The 7-day return was likewise higher (10.7% vs 6.6%; RR 1.62, 95% CI 1.28 to 1.98). The E-value for the 72-hour association was 2.97, meaning an unmeasured confounder would need to be associated with both opioid use and return by a RR of 2.97, beyond the measured covariates, to explain it away.

### Index Admission Was More Than Doubled

First-line opioids were associated with a markedly higher index admission in the full treated cohort (38.1% vs 16.4%; adjusted RR 2.32, 95% CI 2.11 to 2.58; risk difference 21.69 percentage points; E-value 4.08; Figure 3), consistent with substantial confounding by indication and a higher-risk index trajectory among patients selected for opioids; the admission association should be read primarily as evidence of strong baseline severity differences rather than as an independent treatment effect.

### In MIMIC-IV-ED, the 30-Day Return Trajectory Separated Early and Persisted

In MIMIC-IV-ED, in which the by-treatment-class 30-day cumulative-incidence trajectory was constructed, the cumulative incidence of ED return separated by the third day after the index visit and persisted through 30 days: opioid-treated patients accrued returns steadily above the tightly clustered dopamine-antagonist, nonsteroidal, and acetaminophen classes, which were nearly indistinguishable from one another (Figure 2). Intravenous magnesium was too uncommon to characterize (n=128 discharged) and is not plotted in Figure 2; its crude return rates are reported in eTable 6. In crude terms among discharged patients, the 30-day return was 16.5% after opioids versus 11.1% after dopamine antagonists.

**Figure 1.**
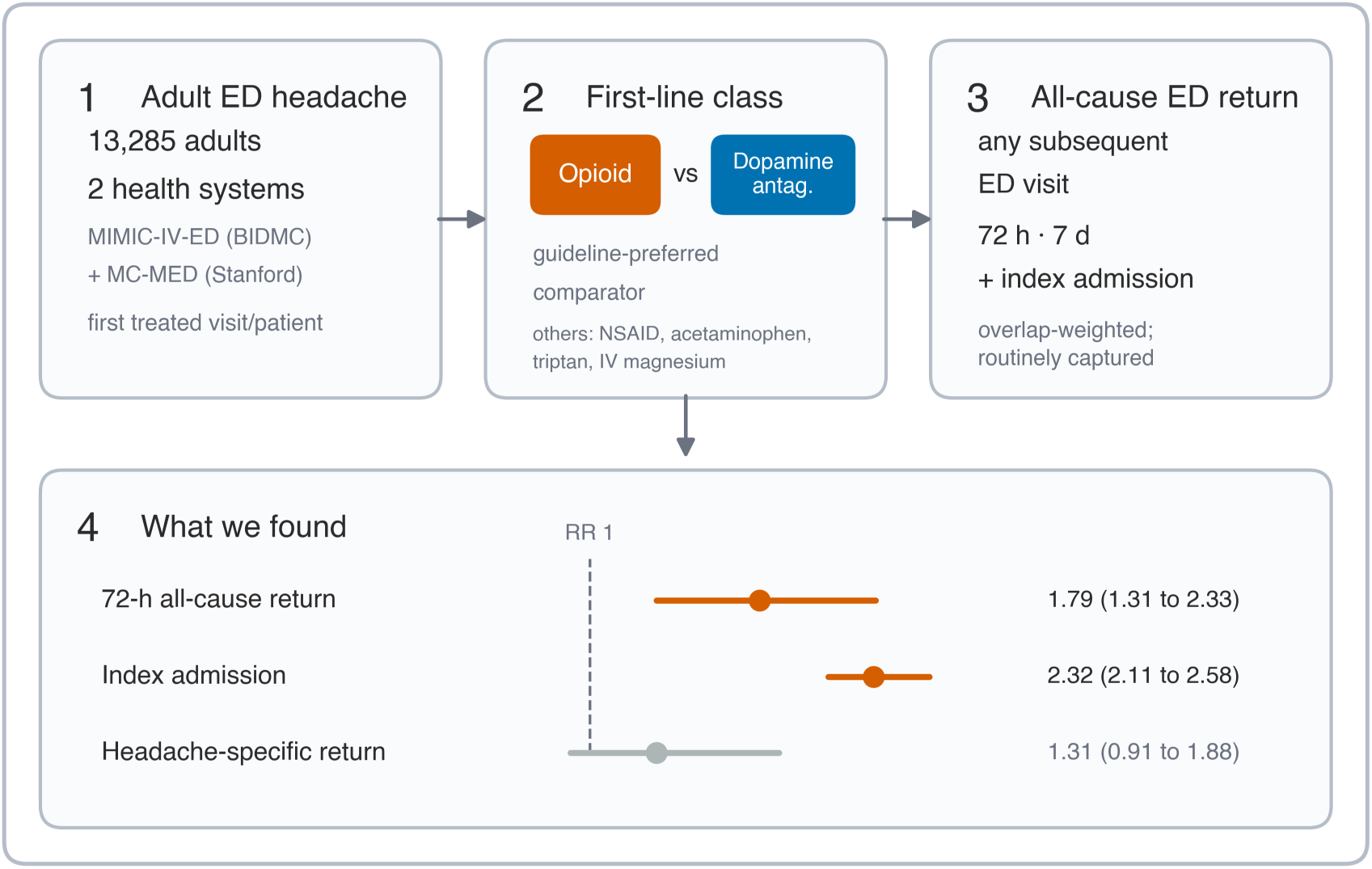
Study overview. Two-center comparative cohort of adults treated for headache in the emergency department (ED). The three input panels show the setting (13,285 adults across MIMIC-IV-ED and MC-MED, first treated visit per patient), the active-comparator contrast (first-line opioid versus guideline-preferred dopamine antagonist, with the other first-line classes listed), and the outcome (all-cause ED return within 72 hours and 7 days and index hospital admission, addressed with propensity overlap weighting). The finding panel shows overlap-weighted risk ratios (opioid versus dopamine antagonist): all-cause return and admission are higher after opioids, whereas the headache-specific return is attenuated and crosses the line of no difference. Index admission is shown as a secondary acute-care outcome and should be interpreted cautiously because it is highly susceptible to confounding by baseline severity. RR, risk ratio.

**Figure 2.**
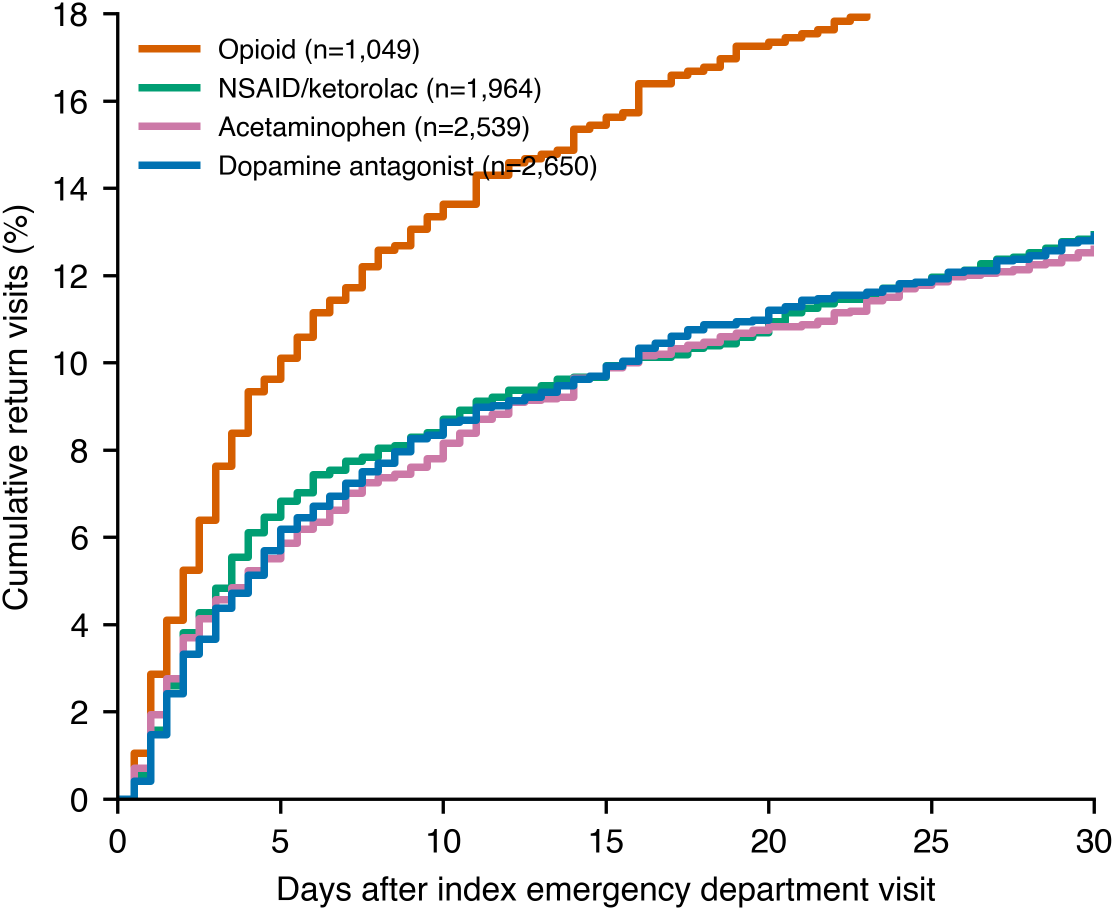
Cumulative incidence of ED return over 30 days by first-line treatment class. Among discharged, treated adult headache patients in the primary cohort (MIMIC-IV-ED), the cumulative percentage with a subsequent ED visit, by first-line class, with in-hospital death as a competing event. The four common first-line classes are shown; the opioid curve separates from the tightly clustered dopamine-antagonist, nonsteroidal, and acetaminophen curves by the third day and remains elevated. The uncommon intravenous-magnesium group (n=128 discharged) is omitted as too small to characterize; its crude rates appear in eTable 6.

### The Direction of Association Was Concordant Across Both Health Systems

The direction of association was concordant across both health systems, although MC-MED return estimates were imprecise (72-hour RR 1.93, 95% CI 0.53 to 4.35) because only 85 opioid-treated discharged patients were available. In MIMIC-IV-ED the adjusted 72-hour return RR was 1.79 (95% CI 1.30 to 2.38) and the admission RR 2.34 (95% CI 2.11 to 2.63). In MC-MED the point estimates were similar or larger (7-day RR 1.95; admission RR 2.23, 95% CI 1.83 to 2.74), with wider confidence intervals reflecting the smaller opioid-treated sample (Figure 3, Figure 4). The pooled two-center estimates, adjusted for site, were RR 1.79 (95% CI 1.31 to 2.33) for the 72-hour return and RR 2.32 (95% CI 2.11 to 2.58) for admission.

**Figure 3.**
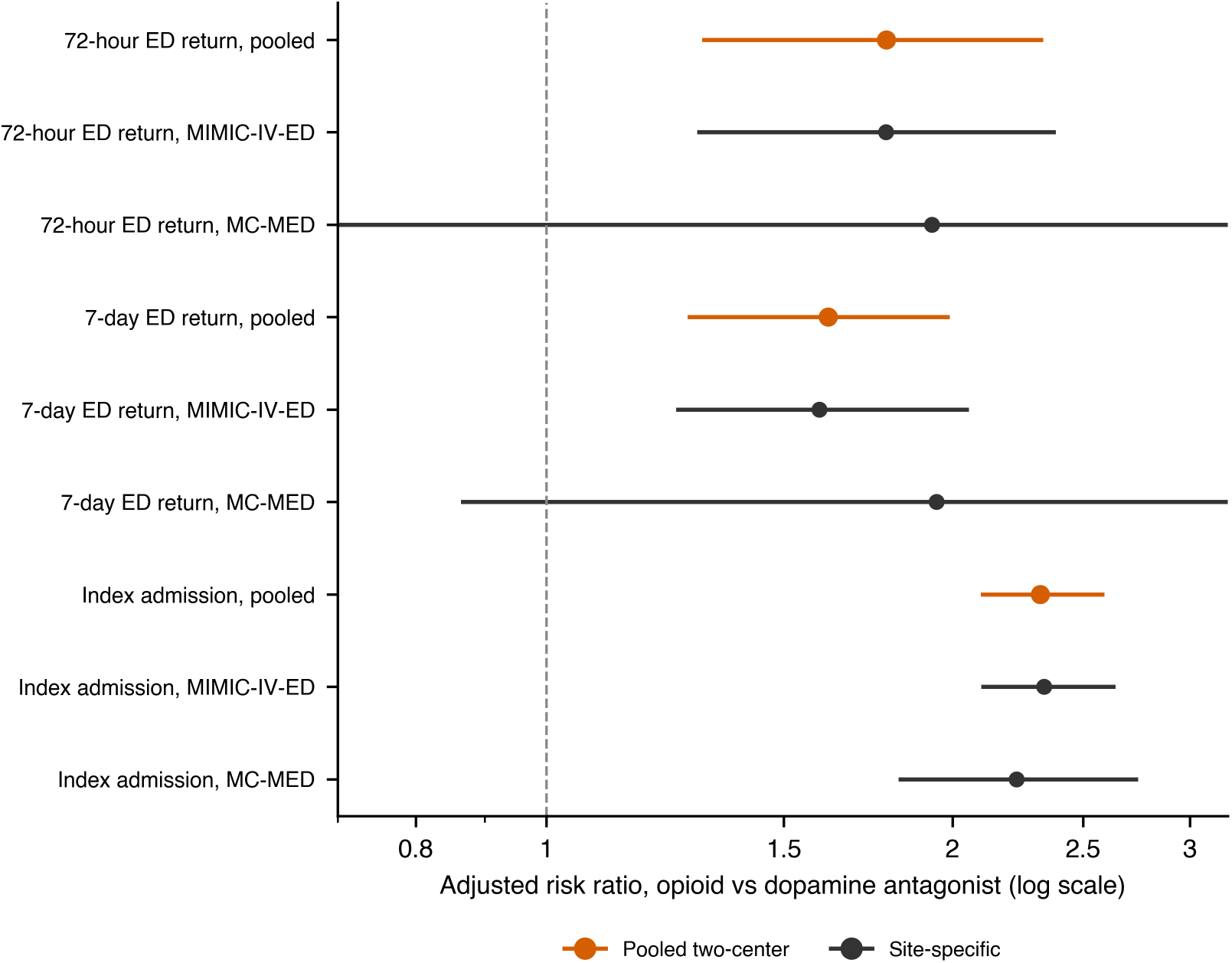
Adjusted risk of return and admission, opioid versus dopamine antagonist. Overlap-weighted risk ratios (opioid versus dopamine antagonist) for the 72-hour ED return, 7-day ED return, and index admission, shown pooled (orange) and by site (MIMIC-IV-ED and MC-MED). Horizontal lines are bootstrap 95% CIs; the dashed line marks no difference. The log-scale x-axis is truncated for display.

**Figure 4.**
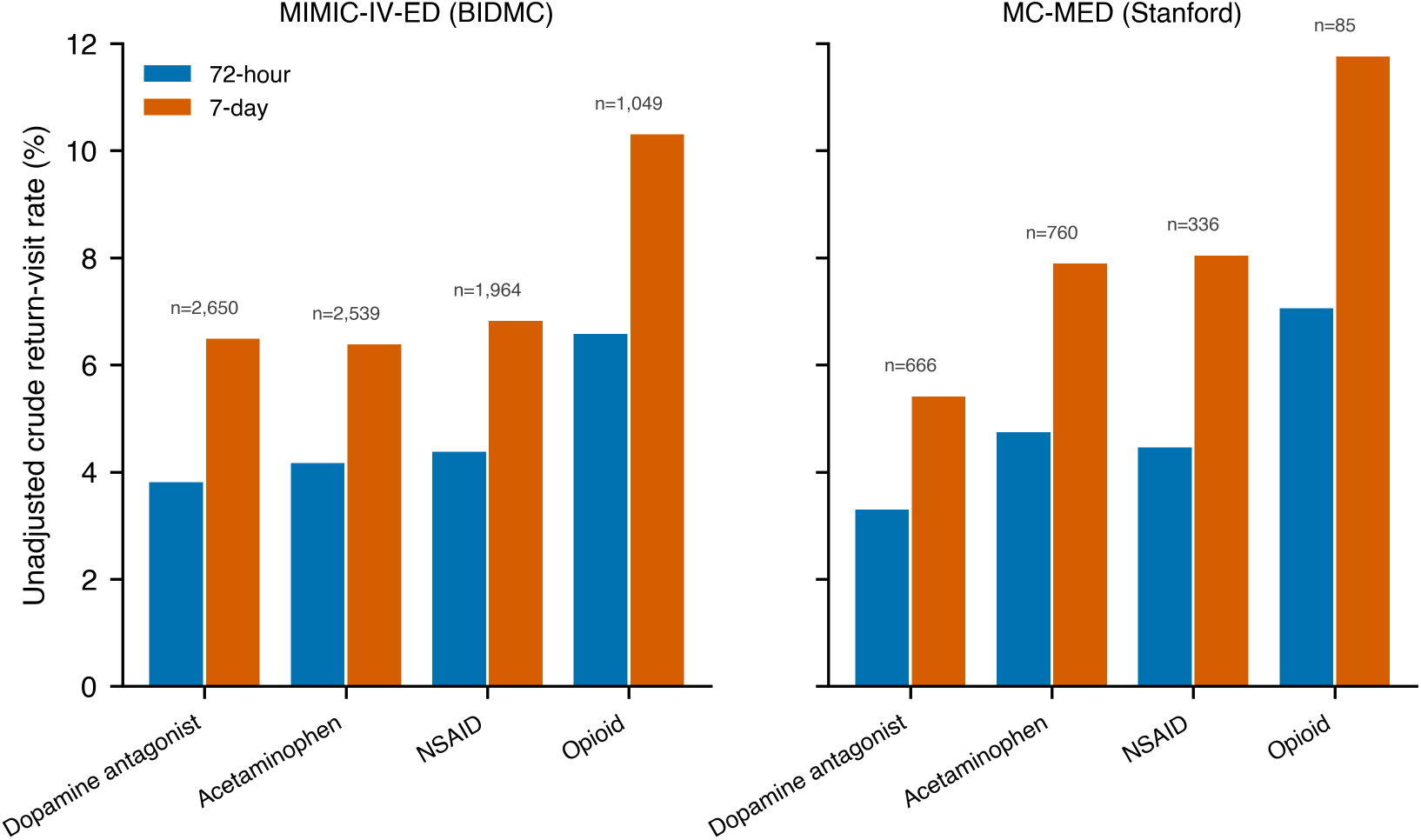
Two-center comparison of crude return by first-line class. Crude 72-hour and 7-day return rates by first-line class in MIMIC-IV-ED (Beth Israel Deaconess) and MC-MED (Stanford), showing the higher opioid return in both health systems.

### Sensitivity Analyses Were Broadly Consistent

After overlap weighting, the effective sample size for the pooled primary 72-hour contrast among discharged patients was 1,078 for opioid recipients and 2,569 for dopamine-antagonist recipients. Excluding index visits with a secondary-headache or red-flag diagnosis removed 115 patients from the MIMIC-IV-ED discharged contrast and left the findings essentially unchanged (72-hour RR 1.77, 95% CI 1.26 to 2.47; 7-day RR 1.63, 95% CI 1.28 to 2.06; admission RR 2.54, 95% CI 2.24 to 2.91). Restricting the dopamine-antagonist comparator to metoclopramide, prochlorperazine, and droperidol gave a similar 72-hour RR of 1.83 (95% CI 1.29 to 2.58), and dropping the 729 visits with a medication tie between opioids and dopamine antagonists gave a comparable 72-hour RR of 1.78 (95% CI 1.19 to 2.59). A composite of index admission or 72-hour return, which did not condition on discharge, was concordant across sites (pooled RR 2.16, 95% CI 1.98 to 2.39; MC-MED RR 2.11, 95% CI 1.74 to 2.58).

Two sensitivity analyses departed from the primary estimate. Restricting to migraine- or primary-headache-coded index visits (601 discharged for the return outcomes) produced higher point estimates for the 7-day return (RR 2.61; 95% CI 1.22 to 6.83) and index admission (RR 4.64; 95% CI 2.07 to 13.22), while the 72-hour estimate was imprecise (RR 2.54; 95% CI 0.83 to 8.83). Because only 102 opioid-treated patients fell in this primary-headache subgroup, these estimates are supportive in direction only and too imprecise to establish a primary-headache-specific effect. When the qualifying return visit itself had to be coded or triaged as headache, the direction of association was preserved but attenuated and no longer statistically significant (pooled 72-hour RR 1.31, 95% CI 0.91 to 1.88; 7-day RR 1.22, 95% CI 0.92 to 1.59), and the MC-MED headache-specific estimate was highly imprecise given its small opioid-treated discharged sample (72-hour RR 1.07; 95% CI 0.00 to 3.75). This attenuation indicates that the all-cause return signal has not been established as headache-specific. Full estimates for each sensitivity analysis are provided in eTables 10 to 14.

## Discussion

In two independent ED cohorts from different health systems, first-line opioid treatment for acute headache was associated with roughly 60% to 80% higher short-term ED return and more than double the index admission compared with guideline-preferred dopamine antagonists. The association held after propensity overlap weighting that closely balanced the prespecified measured covariates available in both databases, was of similar magnitude at both sites, and yielded E-values of 2.97 for the 72-hour return and 4.08 for admission. Because opioid recipients were older and higher-acuity at baseline, the crude difference could have been dismissed as confounding by indication; that it persisted after adjustment and was directionally concordant across both health systems is the central finding.

These results extend the case against routine opioids for ED headache from the in-ED pain endpoint used by randomized trials to a harder, downstream outcome. Trials have shown dopamine antagonists to be more effective than opioids for acute migraine on short-term pain relief, and society guidance recommends against routine opioids.[3,4] Our contribution is that we evaluated a routinely captured downstream utilization outcome, all-cause ED return after an index headache visit, rather than relying only on inconsistently documented in-ED pain reassessment.[7] At scale and across two health systems, first-line opioid treatment was associated with more of this downstream utilization than guideline-preferred dopamine antagonists. The early separation of the return trajectory and the concordance of the three guideline-compatible classes at the low end indicate that this pattern was not shared by the other parenteral or non-opioid classes that were common in this cohort, although intravenous magnesium, the only other parenteral option, was too uncommon to evaluate directly.

Several mechanisms are compatible with these associations and cannot be separated here. Opioids may less effectively abort the underlying headache, may be selected for patients whose headaches are intrinsically more refractory in ways not captured by triage acuity, or may contribute to rebound and medication-overuse patterns that bring patients back. The E-values quantify the strength of association an unmeasured confounder would need with both opioid selection and the outcome to explain away the finding; they do not exclude residual confounding by unmeasured clinical severity, headache subtype, chronic pain or opioid-exposure history, or provider-level practice.

## Limitations

This study has several limitations. First, treatment was not randomized; despite active-comparator design, overlap weighting, and E-values, residual confounding by unmeasured severity is possible, and the findings are associations rather than causal effects. Second, each database observes only returns to its own health system, so out-of-system returns were not captured; this undercounts returns and is expected to be approximately nondifferential across treatment classes and therefore to bias RRs toward the null, although differential out-of-system return by patient complexity or access cannot be excluded. Third, medication exposure was ascertained from dispensing or administration records, which approximate but do not confirm receipt. Fourth, opioid recipients formed a modest fraction of the external MC-MED cohort, so its return estimates were directionally concordant but imprecise, with wider confidence intervals, because the external opioid-treated discharged sample was small. Fifth, headache was identified from chief complaint and diagnosis codes that mix primary and secondary headache; the analysis concerns first-line treatment of undifferentiated ED headache rather than a single diagnosis. Sixth, return outcomes were all-cause ED returns rather than confirmed headache-related revisits; the estimates therefore reflect downstream ED utilization after an index headache visit, not direct measures of headache recurrence or treatment failure. When the outcome was restricted to a headache-specific return, the association was in the same direction but attenuated and no longer statistically significant (pooled 72-hour RR 1.31, 95% CI 0.91 to 1.88), so a headache-specific effect is not established. Seventh, return analyses were conditional on discharge from the index visit, so conditioning on disposition may introduce selection bias; a composite of index admission or 72-hour return, which does not condition on discharge, showed the same direction (pooled RR 2.16, 95% CI 1.98 to 2.39); because this composite includes admission, it shares that outcome’s susceptibility to severity confounding and does not by itself resolve the conditioning. Eighth, several clinically important confounders were unavailable in both databases, including headache subtype, chronic or home opioid use, neuroimaging, focal neurologic deficits, and provider-level practice. The prespecified exclusion and subgroup sensitivity analyses reduce but do not eliminate the concern that first-line opioid use marked unobserved refractory, secondary, chronic-pain, or otherwise high-risk headache presentations.

## Conclusions

Across two health systems, first-line opioid treatment for ED headache was associated with higher short-term all-cause ED return and index admission than guideline-preferred dopamine antagonists, after propensity adjustment and with directionally concordant estimates at both sites. These risk-adjusted associations, measured on an objective downstream outcome rather than in-ED pain, should be interpreted as downstream all-cause ED utilization after an index headache visit rather than proof of headache recurrence or treatment failure; they support opioid-sparing, guideline-concordant first-line treatment and motivate prospective or pragmatic confirmation.

## Funding

None.

## Competing interests

The authors declare that they have no competing interests.

## Data Availability

MIMIC-IV-ED (version 2.2) and MC-MED (version 1.0.0) are available to credentialed users through PhysioNet under the applicable data use agreements. The authors do not redistribute source data.

## Code Availability

Cohort-construction specifications and the numbered analysis scripts that reproduce every reported estimate are available from the corresponding author and will be deposited in a public repository on publication.

## Author Contributions

A.G. conceived the study, performed the analysis, and drafted the manuscript. Y.A. contributed to data engineering. T.L. and O.D. contributed to clinical interpretation. E.K. supervised the work. All authors reviewed and approved the final manuscript.

## Use of AI Tools

Large language models were used only for language editing. All study design, analysis, interpretation, and final content were verified by the authors.

## Notes

### Competing Interest Statement

The authors have declared no competing interest.

