## appendix for "First-Line Opioids and Short-Term All-Cause Emergency Department Return After Headache Visits: A Two-Center Comparative Cohort Study"

This Supplementary Appendix accompanies the main manuscript. It provides the operational cohort definitions, the treatment-class value sets, the full propensity overlap-weighting and E-value procedures, per-site and pooled effect estimates, and covariate-balance diagnostics that support the two-center analysis. All results derive from de-identified data accessed under credentialed PhysioNet data use agreements.

**Contents**

- eMethods. Expanded methods
- eTable 1. Case-identifying diagnosis codes and chief-complaint terms
- eTable 2. First-line treatment classes and constituent agents
- eTable 3. Covariate balance before and after overlap weighting
- eTable 4. Primary and secondary associations, pooled two-center
- eTable 5. Per-site adjusted opioid-versus-dopamine-antagonist estimates
- eTable 6. Crude return-visit rates by first-line class, both databases
- eTable 7. Cohort derivation and first-line class distribution
- eTable 8. E-value sensitivity for the pooled associations
- eTable 9. Covariate balance and effective sample size by analytic population
- eTable 10. Headache-specific return-visit sensitivity analysis

- eTable 11. Composite admission-or-72-hour-return sensitivity analysis
- eTable 12. Migraine/primary-headache-restricted sensitivity analysis
- eTable 13. Secondary-headache and red-flag exclusion sensitivity analysis
- eTable 14. Restricted-comparator and opioid-tie-rule sensitivity analyses
- eTable 15. Observed constituent first-line agents within the opioid and dopamine-antagonist classes
- eTable 16. Raw (unweighted) event counts underlying the primary associations
- eFigure 1. Propensity-score distribution by treatment group
- eFigure 2. Covariate balance (Love plot)


### eMethods. Expanded methods

#### 1. Data sources

The primary cohort was MIMIC-IV-ED (version 2.2), a de-identified database of emergency department (ED) encounters at Beth Israel Deaconess Medical Center between 2011 and 2019. External comparison used MC-MED (version 1.0.0), a de-identified ED database from Stanford University Medical Center spanning approximately 2020 to 2022 on a different electronic health record vendor. The two databases differ in region, calendar period, and clinical documentation system, which provides a strong test of whether any association is an artifact of a single institution. Both are distributed through PhysioNet and were analyzed under the corresponding credentialed data use agreements. In MIMIC-IV-ED, calendar time is available only as de-identified three-year anchor windows, which were used as the calendar-period adjustment term.

### 2. Cohort construction

Adults (age 18 years or older) with an ED visit for headache were identified when the triage chief complaint matched a headache term or when a primary or nonspecific-headache diagnosis code was recorded for the encounter (eTable 1). The treated cohort was restricted to encounters in which at least one qualifying acute agent (eTable 2) was administered. To implement the new-user, active-comparator framing and to avoid within-patient correlation, the unit of analysis was the first treated headache visit per patient; subsequent visits by the same patient contributed only as outcome events. Identical logic was applied in both databases.

### 3. Exposure classification

The first-line treatment class was defined by the earliest qualifying acute agent by administration timestamp. Agents were mapped to seven mutually exclusive classes (eTable 2) by case-insensitive matching on generic and common brand names. When two or more agents shared the earliest timestamp, a tie that included an opioid was classified as opioid. Because opioid co-administration could reflect either ambiguous first-line intent or greater severity, the direction of any resulting bias is uncertain; an alternative tie rule that excluded opioid-containing ties was therefore examined as a sensitivity analysis (eTable 14). The prespecified primary contrast was opioids versus dopamine-receptor antagonists, the guideline-preferred active comparator.

### 4. Outcome ascertainment

The primary outcome was the 72-hour ED return visit, defined as any subsequent ED encounter by the same patient within 72 hours of index ED departure. In MIMIC-IV-ED, return visits were ascertained from linked de-identified subject identifiers and encounter timestamps. In MC-MED, return visits were ascertained from the recorded interval to the next ED encounter. Secondary outcomes were the 7-day and 30-day ED return, index hospital admission, and the cumulative incidence of ED return over 30 days. Because a return visit requires that the patient be discharged from the index visit, all return analyses were restricted to patients discharged at the index encounter; index admission was analyzed in the full treated cohort. In the 30-day cumulative-incidence analysis, in-hospital death was treated as a competing event.

### 5. Covariates

Prespecified confounders available in both databases were age, sex, race and ethnicity, triage acuity, triage pain score, mode of arrival, prior ED utilization, and time of day (categorized as day, evening, or night). The primary database additionally contributed the de-identified calendar-period term, and the pooled model contributed a site indicator. Triage pain score missingness was handled with a missing-indicator term and median substitution; triage acuity missingness was retained as an explicit category.

### 6. Propensity overlap weighting

Confounding by indication was addressed with propensity overlap weights. A logistic-regression propensity model for opioid versus dopamine-antagonist treatment was fit on the standardized covariate design matrix. Each patient received an overlap weight equal to the probability of the opposite treatment: dopamine-antagonist-treated patients were weighted by the estimated probability of opioid treatment, and opioid-treated patients by the estimated probability of dopamine-antagonist treatment. Overlap weighting targets the sub-population with the greatest clinical equipoise and yields exact mean balance on every included covariate, which is not guaranteed by inverse-probability weighting. Balance was summarized by the maximum absolute standardized mean difference (SMD) across all model terms before and after weighting (eTable 3, eFigure 2).

The propensity-score distributions by group are shown in eFigure 1. The same overlap-weighting procedure was repeated independently within each of five additional analytic populations used in the sensitivity analyses below; every population achieved a maximum absolute SMD after weighting of 0.013 or less (eTable 9).

### 7. Effect estimation and uncertainty

Weighted absolute risks were computed as the weighted mean of each binary outcome within treatment group. Risk differences and risk ratios (RRs) were derived from the weighted risks. Confidence intervals were obtained from 1000 patient-level bootstrap resamples in which the propensity model was refit and the weights recomputed within each replicate, so that the reported intervals reflect uncertainty in the weighting as well as in the outcome. The random seed was fixed for reproducibility.

### 8. Sensitivity to unmeasured confounding

Robustness to unmeasured confounding was quantified with the E-value, the minimum strength of association, on the risk-ratio scale, that an unmeasured confounder would need to have with both the treatment and the outcome, conditional on the measured covariates, to explain away the observed association. E-values were computed for each point estimate and for the confidence limit closest to the null (eTable 8).

### 9. Two-center pooling and concordance

Associations were estimated separately in each database and then pooled with a site indicator included in both the propensity and outcome steps. Concordance was assessed by comparing the direction and magnitude of the per-site estimates (eTable 5) and by comparing the crude return-visit ranking of the six first-line classes across databases (eTable 6). MC-MED provided directionally concordant external evidence on a different electronic health record, region, and calendar period.

### 10. Sensitivity analyses

To probe the robustness of the primary opioid-versus-dopamine-antagonist association, six additional analyses were prespecified.

(a) Effective sample size. Because overlap weights are unequal across patients, the weighted analytic sample carries less information than its raw count. Effective sample size was summarized with the Kish formula, $\mathrm{ESS} = (\Sigma \mathrm{w})^2 \ / \ \Sigma \mathrm{w}^2$, computed separately within each treatment arm for six analytic populations: pooled, MIMIC-IV-ED, and MC-MED, each in the discharged 72-hour-return population and in the full treated admission cohort. Covariate balance (maximum absolute SMD) was reconfirmed in each of the six populations (eTable 9).

(b) Headache-specific return. The primary 72-hour and 7-day return outcomes count any subsequent ED visit, regardless of cause. This sensitivity analysis restricted the return event to a subsequent ED visit that was itself coded or triaged as headache, using the same case-identifying criteria as eTable 1, to test whether the primary association reflects headache recurrence specifically rather than any-cause ED return (eTable 10).

(c) Composite admission-or-72-hour-return. Because a return visit requires index discharge, the primary return analyses were restricted to discharged patients. This composite estimand combines index admission and 72-hour return into a single binary outcome evaluated in the full treated cohort, not conditioned on discharge, to summarize the overall short-term acute-care trajectory after the index visit (eTable 11).

(d) Migraine/primary-headache restriction. The primary cohort includes any qualifying headache diagnosis code or chief complaint. This sensitivity analysis restricted the index visit to encounters with a primary migraine or headache-syndrome diagnosis code (ICD-9-CM 346.x, 307.81, 339.x; ICD-10-CM G43, G44), MIMIC-IV-ED only, to exclude visits in which headache was a nonspecific or incidental presenting complaint (eTable 12).

(e) Secondary-headache and red-flag exclusion. Index visits carrying a diagnosis code for a secondary headache etiology or a red-flag condition (intracranial hemorrhage, ischemic stroke or transient ischemic attack, meningitis or encephalitis, CNS neoplasm, head or facial trauma with intracranial injury, hydrocephalus or raised intracranial pressure, temporal arteritis, cerebral venous thrombosis, or eclampsia/pre-eclampsia) were excluded, MIMIC-IV-ED only, to test whether the association persists once visits with an identifiable structural or systemic cause of headache are removed (eTable 13).

(f) Comparator restriction and tie rule. Two further analyses tested definitional choices in the exposure, MIMIC-IV-ED only. The dopamine-antagonist comparator was restricted to the three agents most specific to headache and migraine treatment (metoclopramide, prochlorperazine, droperidol), dropping chlorpromazine and promethazine. Separately, the conservative tie rule that assigned same-timestamp opioid-containing ties to the opioid group (section 3) was reversed by excluding these tied index visits entirely rather than assigning them to opioid (eTable 14).

### 11. Software and reproducibility

Analyses used Python 3.9 with pandas, NumPy, scikit-learn, SciPy, and lifelines. All cohort-construction and analysis scripts are numbered and were run in sequence from raw PhysioNet tables to the reported estimates. No causal effect was estimated; all associations are reported as risk-adjusted.

#### eTable 1. Case-identifying diagnosis codes and chief-complaint terms

An encounter qualified if either a chief-complaint term or a diagnosis code matched. Identical definitions were applied in MIMIC-IV-ED and MC-MED.

| Domain | Criterion |
|---|---|
| Chief-complaint terms (triage free text, case-insensitive) | headache; head ache; migraine; cephalgia; cephalalgia |
| ICD-9-CM diagnosis | 346.x (migraine); 307.81 (tension-type headache); 339.x (other headache syndromes); 784.0 (headache) |
| ICD-10-CM diagnosis | G43 (migraine); G44 (other headache syndromes); R51 (headache) |

#### eTable 2. First-line treatment classes and constituent agents

Agents were matched on generic and common brand names (case-insensitive). Classes are mutually exclusive; the first-line class was the earliest qualifying agent by timestamp. The observed distribution of specific agents within the two contrasted classes is given in eTable 15.

| Class | Constituent agents (match terms) |
|---|---|
| Dopamine-receptor antagonist | metoclopramide, prochlorperazine, droperidol, chlorpromazine, promethazine |
| Opioid | morphine, hydromorphone, oxycodone, hydrocodone, fentanyl, codeine, tramadol, oxymorphone, meperidine, butorphanol, nalbuphine, tapentadol |
| NSAID or ketorolac | ketorolac, ibuprofen, naproxen, ketoprofen, diclofenac, indomethacin, aspirin |
| Triptan | sumatriptan, rizatriptan, zolmitriptan, eletriptan, naratriptan, frovatriptan, almotriptan |
| Dihydroergotamine | dihydroergotamine (DHE-45, dihydroergotamine mesylate) |
| Intravenous magnesium | magnesium sulfate |

| | |
|---|---|
| Acetaminophen | acetaminophen, paracetamol (intravenous or oral) |

### eTable 3. Covariate balance before and after overlap weighting

This table reports the full per-covariate balance detail specifically for the MIMIC-IV-ED discharged primary-contrast population (72-hour-return analysis; n = 3,699; 1,049 opioid-treated). Standardized mean differences (SMD) are for opioid versus dopamine-antagonist treatment; a positive value indicates a higher mean in the opioid group. The de-identified calendar-period terms are summarized by their maximum absolute SMD. Balance and effective sample size for the five other analytic populations (pooled and MC-MED, and the full treated admission cohort) are summarized in eTable 9.

| Covariate | SMD before weighting | SMD after weighting |
|---|---|---|
| Arrival walk-in | -0.349 | 0.000 |
| Triage acuity 3 | -0.338 | 0.000 |
| Age | +0.312 | 0.000 |
| Triage acuity 2 | +0.286 | 0.000 |
| Triage pain score | +0.246 | 0.000 |
| Male sex | +0.170 | 0.000 |
| Triage acuity 4 | +0.128 | 0.000 |
| Triage acuity missing | +0.107 | 0.000 |
| White race | +0.101 | +0.001 |
| Triage pain missing | +0.094 | 0.000 |
| Evening arrival (15-23 h) | +0.079 | 0.000 |
| Black race | -0.072 | 0.000 |
| Prior ED visits | +0.059 | 0.000 |

| Covariate | SMD before weighting | SMD after weighting |
|---|---|---|
| Triage acuity 5 | +0.044 | +0.001 |
| Arrival unknown | -0.027 | -0.001 |
| Arrival other | -0.022 | 0.000 |
| Arrival by helicopter | +0.022 | 0.000 |
| Night arrival (23-7 h) | +0.019 | 0.000 |
| Race other or unknown | +0.005 | 0.000 |
| Hispanic | +0.005 | 0.000 |
| Index calendar period (max across 33 three-year terms) | +0.088 | +0.002 |
| **Maximum absolute SMD (all terms)** | **0.349** | **0.002** |

### eTable 4. Primary and secondary associations, pooled two-center

Overlap-weighted absolute risks, risk differences, and risk ratios for first-line opioids versus dopamine antagonists, pooled across MIMIC-IV-ED and MC-MED with a site term. Return outcomes are among discharged patients; admission is in the full treated cohort. The raw, unweighted event counts (numerators and denominators) underlying these weighted estimates are given in eTable 16. CI, confidence interval; pp, percentage points; RD, risk difference; RR, risk ratio.

| Outcome | Opioid risk, % | Dopamine risk, % | RD, pp (95% CI) | RR (95% CI) | E-value |
|---|---|---|---|---|---|
| 72-hour ED return | 6.77 | 3.79 | +2.98 (1.30 to 4.48) | 1.79 (1.31 to 2.33) | 2.97 |
| 7-day ED return | 10.67 | 6.60 | +4.07 (1.96 to 6.05) | 1.62 (1.28 to 1.98) | 2.62 |
| Index admission | 38.07 | 16.38 | +21.69 (19.19 to 24.29) | 2.32 (2.11 to 2.58) | 4.08 |

### eTable 5. Per-site adjusted opioid-versus-dopamine-antagonist estimates

Overlap-weighted risk ratios by database. Point estimates are in the same direction and of similar or larger magnitude in MC-MED; MC-MED return intervals are wide because only 85 opioid-treated discharged patients were available. CI, confidence interval; RR, risk ratio.

| Outcome | MIMIC-IV-ED RR (95% CI) | MC-MED RR (95% CI) | Pooled RR (95% CI) |
|---|---|---|---|
| 72-hour ED return | 1.79 (1.30 to 2.38) | 1.93 (0.53 to 4.35) | 1.79 (1.31 to 2.33) |
| 7-day ED return | 1.59 (1.25 to 2.05) | 1.95 (0.87 to 3.93) | 1.62 (1.28 to 1.98) |
| Index admission | 2.34 (2.11 to 2.63) | 2.23 (1.83 to 2.74) | 2.32 (2.11 to 2.58) |

### eTable 6. Crude return-visit rates by first-line class, both databases

Unadjusted return-visit rates among discharged patients by first-line class. The ranking is concordant across databases: opioids carry the highest short-term return in both, and dopamine antagonists, NSAIDs, and acetaminophen cluster together at a lower level. The by-treatment-class 30-day breakdown was computed only for MIMIC-IV-ED; MC-MED's overall 30-day return rate (16.4%, all treated classes combined) does exist and is reported in the main manuscript Results. Triptans and magnesium were uncommon and are shown for completeness.

| First-line class | MIMIC n | MIMIC 72 h, % | MIMIC 7 d, % | MIMIC 30 d, % | MC-MED n | MC-MED 72 h, % | MC-MED 7 d, % |
|---|---|---|---|---|---|---|---|
| Opioid | 1,049 | 6.58 | 10.30 | 16.49 | 85 | 7.06 | 11.76 |
| Dopamine antagonist | 2,650 | 3.81 | 6.49 | 11.06 | 666 | 3.30 | 5.41 |
| NSAID or ketorolac | 1,964 | 4.38 | 6.82 | 11.25 | 336 | 4.46 | 8.04 |
| Acetaminophen | 2,539 | 4.17 | 6.38 | 11.26 | 760 | 4.74 | 7.89 |
| Intravenous magnesium | 128 | 8.59 | 10.16 | 17.97 | 37 | 5.41 | 8.11 |

| First-line class | MIMIC n | MIMIC 72 h, % | MIMIC 7 d, % | MIMIC 30 d, % | MC-MED n | MC-MED 72 h, % | MC-MED 7 d, % |
|---|---|---|---|---|---|---|---|
| Triptan | 16 | 6.25 | 6.25 | 18.75 | 7 | 0.00 | 14.29 |

### eTable 7. Cohort derivation and first-line class distribution

Derivation of the analytic cohorts and the distribution of first-line treatment class in each database. Percentages are of the treated analytic cohort. Dihydroergotamine was included in the treatment-class definition but was not observed as a first-line agent in either database.

| Step | MIMIC-IV-ED | MC-MED |
|---|---|---|
| ED encounters in source database | 425,087 | 118,385 |
| Headache encounters (code or chief complaint) | 19,513 | 5,620 |
| Treated with a qualifying acute agent | 13,693 | 2,788 |
| First treated visit per patient (analytic cohort) | 10,799 | 2,486 |
| First-line dopamine antagonist | 3,056 (28.3%) | 815 (32.8%) |
| First-line opioid | 1,895 (17.5%) | 172 (6.9%) |
| First-line NSAID or ketorolac | 2,304 (21.3%) | 399 (16.1%) |
| First-line acetaminophen | 3,361 (31.1%) | 1,038 (41.8%) |
| First-line intravenous magnesium | 167 (1.5%) | 55 (2.2%) |
| First-line triptan | 16 (0.1%) | 7 (0.3%) |

### eTable 8. E-value sensitivity for the pooled associations

E-value for each pooled point estimate and for the confidence limit closest to the null. The point-estimate E-values near 3 for the return outcomes indicate that an unmeasured confounder would need a risk-ratio association of roughly 3 with both opioid treatment and ED return, beyond the balanced covariates, to fully account for the finding. CI, confidence interval; RR, risk ratio.

| Association | Pooled RR (95% CI) | E-value, point estimate | E-value, CI limit |
|---|---|---|---|
| 72-hour ED return | 1.79 (1.31 to 2.33) | 2.97 | 1.95 |
| 7-day ED return | 1.62 (1.28 to 1.98) | 2.62 | 1.88 |
| Index admission | 2.32 (2.11 to 2.58) | 4.08 | 3.64 |

### eTable 9. Covariate balance and effective sample size by analytic population

Maximum absolute standardized mean difference (SMD) before and after overlap weighting, and Kish effective sample size (ESS) by treatment arm, for each of the six analytic populations used in the manuscript and this appendix; every population was balanced to a maximum absolute SMD after weighting of 0.013 or less.

| Analytic population | n | Opioid n | Max SMD before | Max SMD after | Effective n, opioid | Effective n, dopamine |
|---|---|---|---|---|---|---|
| Pooled, discharged (72h return) | 4,450 | 1,134 | 0.371 | 0.003 | 1,078 | 2,569 |
| MIMIC-IV-ED, discharged (72h return) | 3,699 | 1,049 | 0.349 | 0.001 | 998 | 2,196 |
| MC-MED, discharged (72h return) | 751 | 85 | 0.305 | 0.013 | 84 | 498 |
| Pooled, full cohort (admission) | 5,938 | 2,067 | 0.481 | 0.001 | 1,840 | 2,998 |
| MIMIC-IV-ED, full cohort (admission) | 4,951 | 1,895 | 0.509 | 0.001 | 1,684 | 2,488 |
| MC-MED, full cohort (admission) | 987 | 172 | 0.323 | 0.009 | 167 | 625 |

Values were recomputed with the shared overlap-weighting estimator; small differences from eTable

3 in the third decimal reflect independent propensity-model fits and are not material.

### eTable 10. Headache-specific return-visit sensitivity analysis

Overlap-weighted risk ratios when the return event is restricted to a subsequent ED visit that is itself coded or triaged as headache (eMethods, section 10b), pooled and by site; the pooled 72-hour association attenuates to a risk ratio of 1.31, with a confidence interval that now includes the null, indicating that part of the primary all-cause return signal reflects returns for reasons other than recurrent headache. CI, confidence interval; RR, risk ratio.

| Population | Outcome | n | Opioid n | RR (95% CI) | E-value |
|---|---|---|---|---|---|
| Pooled | Headache-coded 72-hour return | 4,450 | 1,134 | 1.31 (0.91 to 1.88) | 1.95 |
| Pooled | Headache-coded 7-day return | 4,450 | 1,134 | 1.22 (0.92 to 1.59) | 1.74 |
| MIMIC-IV-ED | Headache-coded 72-hour return | 3,699 | 1,049 | 1.34 (0.90 to 1.92) | 2.02 |
| MIMIC-IV-ED | Headache-coded 7-day return | 3,699 | 1,049 | 1.24 (0.90 to 1.64) | 1.78 |
| MC-MED | Headache-coded 72-hour return | 751 | 85 | 1.07 (0.00 to 3.75)* | 1.33 |
| MC-MED | Headache-coded 7-day return | 751 | 85 | 1.08 (0.00 to 2.77)* | 1.37 |

*The two MC-MED headache-specific estimates are highly imprecise. The small opioid-treated discharged sample (85 patients) contained no headache-coded returns in a substantial share of bootstrap resamples, so the 2.5th percentile of the resampled risk ratio reached zero. Point estimates are shown for completeness but are uninformative.

### eTable 11. Composite admission-or-72-hour-return sensitivity analysis

Overlap-weighted risk ratios for the composite outcome of index admission or 72-hour ED return, evaluated in the full treated cohort without conditioning on index discharge (eMethods, section 10c), pooled and by site; the composite association is larger than the return-only estimate because it also captures the strong admission effect. CI, confidence interval; RR, risk ratio.

| Population | n | Opioid n | RR (95% CI) | E-value |
|---|---|---|---|---|
| Pooled | 5,938 | 2,067 | 2.16 (1.98 to 2.39) | 3.74 |
| MIMIC-IV-ED | 4,951 | 1,895 | 2.17 (1.96 to 2.40) | 3.76 |
| MC-MED | 987 | 172 | 2.11 (1.74 to 2.58) | 3.63 |

#### eTable 12. Migraine/primary-headache-restricted sensitivity analysis

Overlap-weighted risk ratios when the index visit is restricted to a primary migraine or headache-syndrome diagnosis code (ICD-9-CM 346.x, 307.81, 339.x; ICD-10-CM G43, G44; eMethods, section 10d), MIMIC-IV-ED only; point estimates are larger than in the full cohort, though the smaller restricted sample widens the 72-hour confidence interval to include the null. CI, confidence interval; RR, risk ratio.

| Outcome | n | Opioid n | RR (95% CI) | E-value |
|---|---|---|---|---|
| 72-hour ED return | 601 | 102 | 2.54 (0.83 to 8.83) | 4.51 |
| 7-day ED return | 601 | 102 | 2.61 (1.22 to 6.83) | 4.65 |
| Index admission | 636 | 118 | 4.64 (2.07 to 13.22) | 8.76 |

#### eTable 13. Secondary-headache and red-flag exclusion sensitivity analysis

Overlap-weighted risk ratios after excluding index visits carrying a diagnosis code for a secondary headache etiology or red-flag condition (eMethods, section 10e), MIMIC-IV-ED only. The exclusion set comprised 19,054 ED encounters in the source database that carried at least one qualifying secondary/red-flag diagnosis code; the headache cohort was rebuilt after removing any overlap with this set. The return-visit associations were essentially unchanged after this exclusion (72-hour and

7-day risk ratios comparable to the primary analysis), while the index-admission association was similar though slightly higher. CI, confidence interval; RR, risk ratio.

| Outcome | n | Opioid n | RR (95% CI) | E-value |
|---|---|---|---|---|
| 72-hour ED return | 3,584 | 979 | 1.77 (1.26 to 2.47) | 2.94 |
| 7-day ED return | 3,584 | 979 | 1.63 (1.28 to 2.06) | 2.65 |
| Index admission | 4,593 | 1,638 | 2.54 (2.24 to 2.91) | 4.52 |

#### eTable 14. Restricted-comparator and opioid-tie-rule sensitivity analyses

Two definitional sensitivity analyses, MIMIC-IV-ED only (eMethods, section 10f): restricting the dopamine-antagonist comparator to metoclopramide, prochlorperazine, and droperidol (2,500 index visits met this narrower definition, dropping chlorpromazine and promethazine), and excluding the 729 index visits whose earliest medication administration was an opioid tied at the same timestamp with another agent, rather than assigning them to the opioid group as under the primary tie rule. Both restrictions preserve the direction and approximate magnitude of the primary associations. CI, confidence interval; RR, risk ratio.

| Analysis | Outcome | n | Opioid n | RR (95% CI) | E-value |
|---|---|---|---|---|---|
| Restricted comparator | 72-hour ED return | 3,164 | 1,049 | 1.83 (1.29 to 2.58) | 3.06 |
| Restricted comparator | 7-day ED return | 3,164 | 1,049 | 1.69 (1.30 to 2.22) | 2.77 |
| Restricted comparator | Index admission | 4,311 | 1,895 | 2.52 (2.26 to 2.87) | 4.47 |
| Drop opioid ties | 72-hour ED return | 3,267 | 617 | 1.78 (1.19 to 2.59) | 2.95 |
| Drop opioid ties | 7-day ED return | 3,267 | 617 | 1.68 (1.28 to 2.18) | 2.75 |
| Drop opioid ties | Index admission | 4,222 | 1,166 | 2.35 (2.09 to 2.66) | 4.13 |

### eTable 15. Observed constituent first-line agents within the opioid and dopamine-antagonist classes

The specific agent administered as the earliest qualifying medication, tabulated within the two contrasted first-line classes for each database. Counts are for the analytic cohort (the first treated headache visit per patient) and reconcile to the class totals in eTable 7. The opioid class comprised predominantly parenteral short-acting opioids rather than weak oral agents (tramadol accounted for 3.0% of opioid-treated patients in MIMIC-IV-ED and none in MC-MED), and the dopamine-antagonist class comprised predominantly the guideline-preferred antiemetics metoclopramide and prochlorperazine; promethazine, a less headache-specific agent, accounted for under 2% of the class in either database. Percentages are within class and database and may not sum to exactly 100 because of rounding.

| Database | Class | Agent | n | % of class |
|---|---|---|---|---|
| MIMIC-IV-ED | Opioid | Morphine | 939 | 49.6 |
| | | Oxycodone | 437 | 23.1 |
| | | Hydromorphone | 339 | 17.9 |
| | | Fentanyl | 84 | 4.4 |
| | | Tramadol | 56 | 3.0 |
| | | Hydrocodone | 33 | 1.7 |
| | | Codeine | 7 | 0.4 |
| | | **Opioid, all agents** | **1,895** | **100.0** |
| | Dopamine antagonist | Metoclopramide | 2,223 | 72.7 |
| | | Prochlorperazine | 773 | 25.3 |
| | | Promethazine | 48 | 1.6 |
| | | Droperidol | 8 | 0.3 |
| | | Chlorpromazine | 4 | 0.1 |

| Database | Class | Agent | n | % of class |
|---|---|---|---|---|
| | | **Dopamine antagonist, all agents** | **3,056** | **100.0** |
| MC-MED | Opioid | Hydromorphone | 81 | 47.1 |
| | | Morphine | 42 | 24.4 |
| | | Hydrocodone | 21 | 12.2 |
| | | Oxycodone | 18 | 10.5 |
| | | Fentanyl | 10 | 5.8 |
| | | **Opioid, all agents** | **172** | **100.0** |
| | Dopamine antagonist | Metoclopramide | 365 | 44.8 |
| | | Prochlorperazine | 361 | 44.3 |
| | | Droperidol | 84 | 10.3 |
| | | Promethazine | 5 | 0.6 |
| | | **Dopamine antagonist, all agents** | **815** | **100.0** |

### eTable 16. Raw (unweighted) event counts underlying the primary associations

Numerators and denominators for each primary outcome by first-line class, shown as events/patients (crude percentage), before any weighting. Return outcomes are among discharged patients; index admission is in the full treated cohort. These crude counts are unweighted and therefore differ from the overlap-weighted risks in eTable 4, which target the clinical-equipoise subpopulation; the crude percentages reconcile with the by-class rates in eTable 6. Pooled rows sum the two databases.

| Outcome | Database | Opioid, events/n (%) | Dopamine antagonist, events/n (%) |
|---|---|---|---|
| 72-hour ED return (discharged) | MIMIC-IV-ED | 69/1,049 (6.58) | 101/2,650 (3.81) |
| | MC-MED | 6/85 (7.06) | 22/666 (3.30) |

| Outcome | Database | Opioid, events/n (%) | Dopamine antagonist, events/n (%) |
|---|---|---|---|
| | Pooled | 75/1,134 (6.61) | 123/3,316 (3.71) |
| 7-day ED return (discharged) | MIMIC-IV-ED | 108/1,049 (10.30) | 172/2,650 (6.49) |
| | MC-MED | 10/85 (11.76) | 36/666 (5.41) |
| | Pooled | 118/1,134 (10.41) | 208/3,316 (6.27) |
| Index admission (full cohort) | MIMIC-IV-ED | 797/1,895 (42.06) | 348/3,056 (11.39) |
| | MC-MED | 87/172 (50.58) | 149/815 (18.28) |
| | Pooled | 884/2,067 (42.77) | 497/3,871 (12.84) |

### eFigure legends

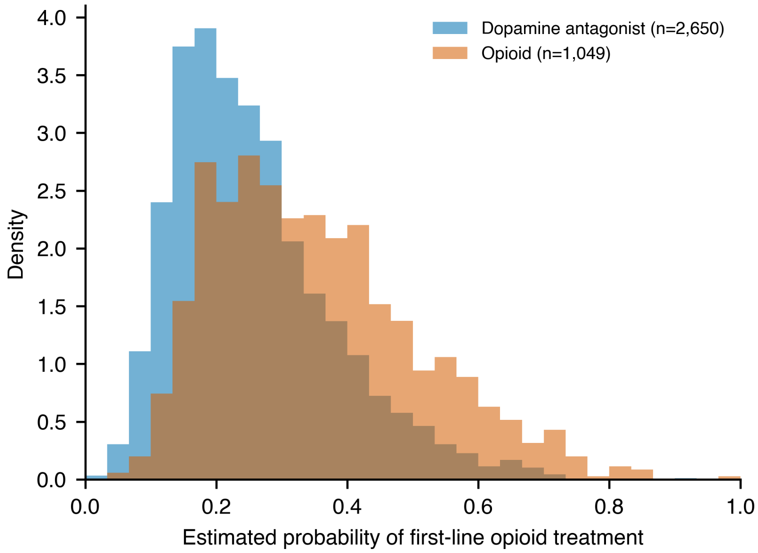


**eFigure 1. Propensity-score distribution by treatment group.** Estimated probability of first-line opioid treatment from the propensity model, shown separately for dopamine-antagonist-treated and opioid-treated patients in the MIMIC-IV-ED discharged primary-contrast population. The broad region of common support indicates that the two groups overlap across most of the

propensity range, the condition under which overlap weighting is well behaved.

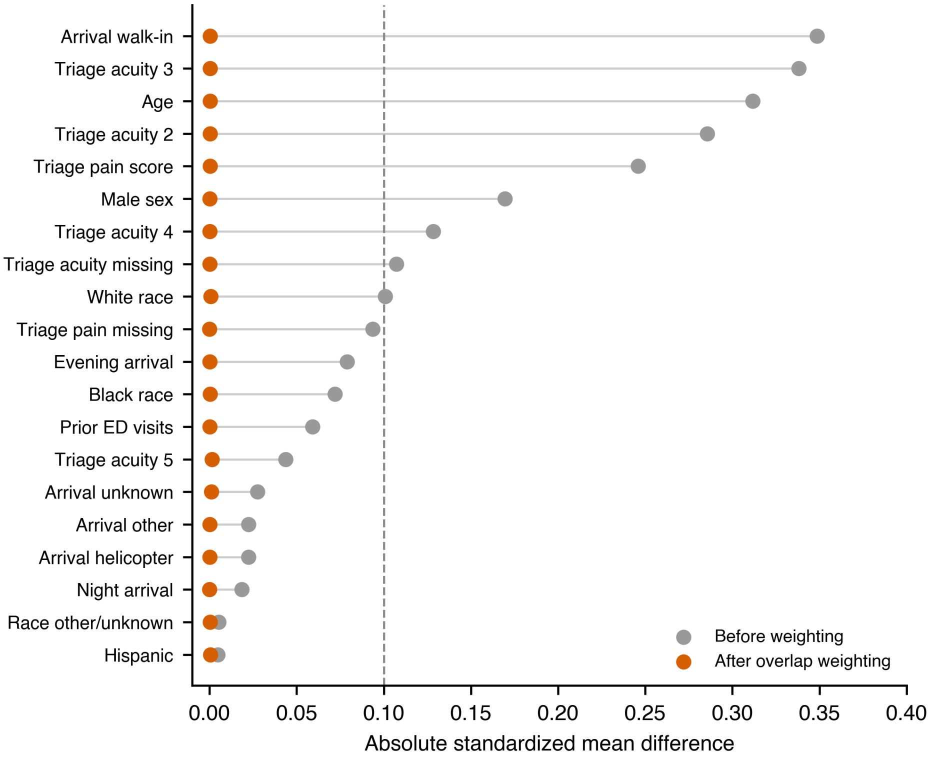


**eFigure 2. Covariate balance (Love plot).** Absolute standardized mean difference for each covariate before (grey) and after (orange) overlap weighting, MIMIC-IV-ED discharged primary-contrast population. The dashed line marks the conventional 0.1 threshold. Before weighting, several severity-related covariates exceed the threshold; after weighting, all covariates are balanced to within 0.002.

### Code and data availability

MIMIC-IV-ED (version 2.2) and MC-MED (version 1.0.0) are available to credentialed users through PhysioNet under their respective data use agreements. The numbered cohort-construction and analysis scripts that reproduce every number in the manuscript and this appendix are available from the corresponding author and will be deposited in a public repository on acceptance. No individual-level data are redistributed.
